# *ApoE* e2 and aging-related outcomes in 379,000 UK Biobank participants

**DOI:** 10.1101/2020.02.12.20022459

**Authors:** Chia-Ling Kuo, Luke C. Pilling, Janice L. Atkins, George A. Kuchel, David Melzer

## Abstract

The *Apolipoprotein E (APOE)* e4 allele is associated with reduced longevity and increased Coronary Artery Disease (CAD) and Alzheimer’s disease, with e4e4 having markedly larger effect sizes than e3e4. The e2 longevity promoting variant is less studied. We conducted a phenome-wide association study of *ApoE* e2e3 and e2e2 with aging phenotypes, to assess their potential as targets for anti-aging interventions. Data were from 379,000 UK Biobank participants, aged 40 to 70 years. e2e3 (n=46,535) had mostly lower lipid-related biomarker levels including reduced total and LDL-cholesterol, and lower risks of CAD (Odds Ratio=0.87, 95% CI: 0.83 to 0.90, p=4.92×10^−14^) and hypertension(OR=0.94, 95% CI: 0.92 to 0.97, p=7.28×10^−7^) versus e3e3. However, lipid changes in e2e2 (n=2,398) were more extreme, including a marked increase in triglyceride levels (0.41 Standard Deviations, 95% CI: 0.37 to 0.45, p=5.42×10^−92^), with no associated changes in CAD risks. There were no associations with biomarkers of kidney function. The effects of both e2e2 and e2e3 were minimal on falls, muscle mass, grip strength or frailty. In conclusion, e2e3 has protective effects on some health outcomes, but the effects of e2e2 are not similar, complicating the potential usefulness of e2 as a target for anti-aging intervention.

## Introduction

The *APOE (Apolipoprotein E)* gene has three major isoforms named APOE2, APOE3, and APOE4. These isoforms are coded by e2, e3, and e4 alleles, which are the haplotypes of the single nucleotide polymorphisms (SNPs), rs429358 and rs7412 on chromosome 19 (T-T, C-T, and C-C, respectively). e2 is associated with decreasing apolipoprotein E, followed by e3 and then e4. The allele-specific, isoform difference gives the variation in domain interaction, protein stability, and protein folding, which influence various pathologies [1][2], where APOE plays the role of shuttling cholesterol and other lipids between cells in the periphery and the central nervous system [3].

In European-ancestry populations, the frequencies of e2, e3, and e4 are approximately 8%, 78%, and 13%, similarly in men and in women [4]. The majority of the population are e3e3 homozygotes (63%), followed by e4e3 (19%) and then e2e3 (13%) [4]. While the e3 allele is the most abundant allele, e4 is the ancestral allele and e2 emerged after e3 [5]. The e2 allele is the youngest (8,000 years ago from east Asia) but under positive selection, expected to have strong evolutionary advantages [5].

Previous studies have shown that the risk of Alzheimer’s disease in e4e4 homozygotes (OR=14.9, 95% CI=10.8 to 20.6) is more than double the risk of e4e3 (OR=3.2, 95% CI=2.8 to 3.8) [6]. Assuming a similar pattern, we hypothesized that the effect of e2e2 is much stronger than that of e2e3 on aging phenotypes and we aimed to characterize individual genotypic effects. e2 has been associated with longevity and reduced risks of Alzheimer’s disease, dementia with Lewy bodies, and cardiovascular diseases (coronary artery disease, myocardial infarction (MI), ischemic stroke). e2 has also been associated with increased high-density lipoprotein cholesterol (HDL), and decreased total cholesterol and low-density lipoprotein (LDL) cholesterol [1][3][7]. The association between e2 and longevity has been robustly replicated across studies [3][7][8][9] and may be partly attributed to negative associations with diseases and conditions. However, e2 has been linked to diseases including age-related macular degeneration (AMD) [10][11], renal disease [12], lipid metabolism disorders (type III hyperlipidemia, high triglycerides or hypertriglyceridemia), and cerebrovascular diseases (cerebral amyloid angiopathy that frequently causes lobar hemorrhagic stroke) [3]. All of these detrimental effects need to be considered to leverage the efficacy of e2-based therapeutics.

In general, drug targets with genetic evidence support are more likely to succeed in human trials [13]. Moreover, associations between e2 and multiple aging traits including longevity suggest that if the underlying shared aging pathways were to be targeted, such an approach may delay the onset of multiple diseases, consistent with the geroscience hypothesis [14]. To characterize e2 in aging, we conducted a phenome-wide association study to associate *ApoE* genotypes to a variety of aging traits in UK Biobank, with the focus on e2e2 and e2e3. The UK Biobank is well-suited to this analysis as it includes thousands of e2e2s and e2e3s and a wealth of baseline measures plus updated mortality and disease diagnoses.

## Results

Of 379,703 unrelated, European-descent participants (Table 1), 54% were women (n=204,726). The mean age at recruitment was 56.7 years (SD=8.0). Participants were followed to death or the last update of survival (Feb 15, 2018), with the mean follow-up time of 9.4 years (SD=1.2). During follow-up, 15,439 participants died and the mean age at death was 67.3 years (SD=7.0). The *ApoE* genotype distribution was similar to that in the general white population. 2,398 participants were e2e2 homozygotes and the sample size of e2e3 was 46,525. e1e2 and e1e4 (n=18 in total) were too few to study; thus, were excluded from analyses. A summary of studied aging associated biomarkers, diagnoses, plus chronic pain, functional measures and frailty (here termed ‘aging traits’) for separate *ApoE* genotypes is provided in Supplementary Table S1.

**Table 1.** Included Participant Characteristics.

| Variable | Mean ± SD or Frequency (%) |
| --- | --- |
| Sex (=female) | 204,736 (54%) |
| Age at recruitment (years) | 56.7 ± 8.0 |
| Follow-up time (years) | 9.4 ± 1.2 |
| Death status at Feb, 2018 | 15,439 (4.1%) |
| Age at Death | 67.3 ± 7.0 |
| *ApoE* genotype |  |
| e1e2 | 3 (<0.01%) |
| e1e4 | 15 (<0.01%) |
| e2e2 | 2,398 (0.63%) |
| e2e3 | 46,535 (12.26%) |
| e2e4 | 9,490 (2.50%) |
| e3e3 | 222,225 (58.53%) |
| e3e4 | 90,016 (23.71%) |
| e4e4 | 9,021 (2.38%) |

The focus of this study is on e2e3 and e2e2; however, all the associations between *ApoE* genotypes and aging traits including those with e4 genotypes are presented in Supplementary Table S2.

### Biomarkers

In Figure 1, we highlight the associations with biomarkers that reached the Bonferroni-adjusted significance level of 5% and showed a 0.1 SD or larger mean difference between e2e2 or e2e3 and e3e3. Compared to e3e3, e2e2 tended to be more associated than e2e3 with biomarkers associated with CAD. Both had lower mean total cholesterol (−0.78 SD in e2e2, p=1.56×10^−314^ versus −0.33 SD in e2e3, p<1×10^−323^), LDL cholesterol (−1.12 SD in e2e2, p<1×10^−323^ versus −0.43 SD in e2e3, p<1×10^−323^), lipoprotein A (−0.40 SD in e2e2, p=4.58×10^−62^ versus − 0.10 SD in e2e3, p=1.41×10^−61^), and apolipoprotein B (−1.99 SD in e2e2, p<1×10^−323^ versus −0.56 SD in e2e3, p<1×10^−323^), plus higher apolipoprotein A1 (0.13 SD in e2e2, p=8.37×10^−11^ versus 0.11 SD in e2e3, p=1.51×10^−104^), all associated with lower risks of CAD. e2e2 and e2e3, however, had higher mean triglycerides (0.41 SD in e2e2, p=5.42×10^−92^ versus 0.11 SD in e2e3, p=1.04×10^−97^), which is associated with higher risks of CAD.

**Figure 1.**
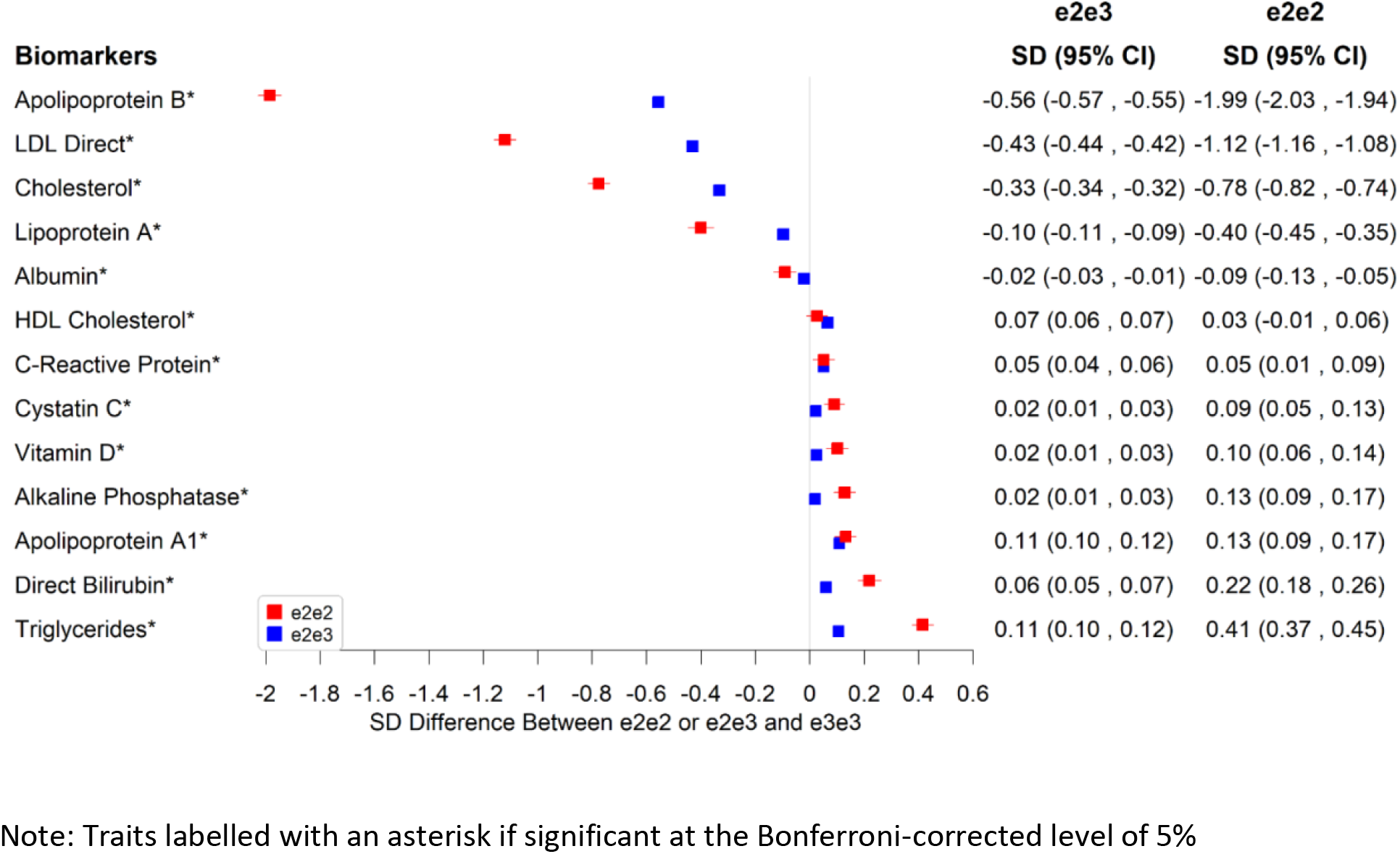
Significant Associations Between e2e2 or e2e3 and Biomarkers at the Bonferroni-Corrected level of 5%.

e2e2 and e2e3 were associated with lower albumin and higher direct bilirubin and alkaline phosphatase, but the effect of e2e3 was not as striking as that of e2e2, e.g., 0.22 SD higher in e2e2 (p=5.12×10^−24^) versus 0.06 SD higher in e2e3 compared to e3e3 for direct bilirubin (p=3.20×10^−28^). Additionally, the mean vitamin level of e2e2 was higher than that of e3e3 by 0.1 SD. e3e3 and other *ApoE* genotypes shared similar vitamin D levels.

There were also associations with various hematology measures (Figure 2 and Supplementary Table S2), including on reticulocyte numbers, with e2e2 being associated with much larger effects than of e2e3. Compared to e3e3, e2e2 had 0.22 SD lower mean reticulocyte count (p=7.90×10^−27^), 0.30 SD lower high light scatter reticulocyte count (p=3.78×10^−47^), 0.31 SD lower immature reticulocyte fraction (p=3.07×10^−50^), and 0.09 SD higher mean reticulocyte volume (p=8.46×10^−6^). e2e2 also had 0.43 SD higher red cell distribution width (RDW) than e3e3 (p=2.16×10), 0.14 SD lower mean corpuscular hemoglobin (p=5.84×10^−12^), 0.12 SD lower mean corpuscular volume (p=1.51×10^−9^), 0.11 SD higher mean platelet volume (p=4.97×10^−8^), and 0.13 SD higher mean sphered cell volume (p=2.12×10^−10^).

**Figure 2.**
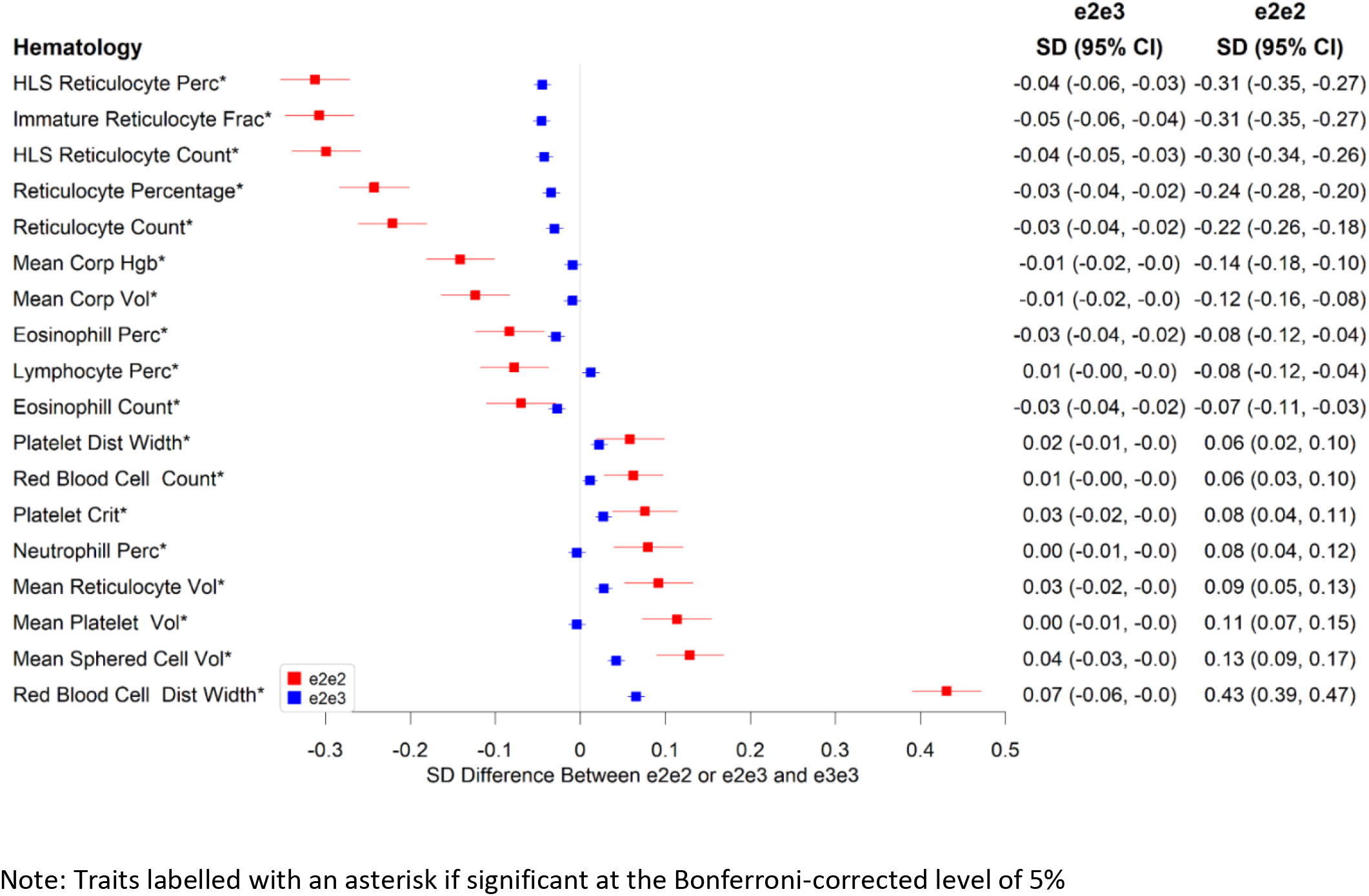
Significant Associations Between e2e2 or e2e3 and Hematological Measures at the Bonferroni-Corrected level of 5%.

### Disease Outcomes

The results for diseases with ~80% power or higher to detect odds ratios approximately 1.2 and 1.2^2^ comparing e2e3, and e2e2 to e3e3 are presented in Figure 3. All the disease association results can be found in Supplementary Table S2. CAD and hypertension were the two significant diseases with e2e2 and/or e2e2 effect compared to e3e3 at the 5% Bonferroni-adjusted level. e2e3 had a lower risk of hypertension than e3e3 (OR=0.94, 95% CI: 0.92 to 0.97, p=7.28×10^−7^) but the protective effect was reduced in e2e2 (OR=1.01, 95% CI: 0.92 to 1.10, p=0.911). Similarly, e2e3 heterozygotes were protected from CAD but the association was not seen with e2e2: the odds ratio for CAD was 0.87 (95% CI: 0.83 to 0.90, p=4.92×10^−14^) comparing e2e3 to e3e3 and was 1.04 (95% CI: 0.90 to 1.19, p=0.635) comparing e2e2 to e3e3.

**Figure 3.**
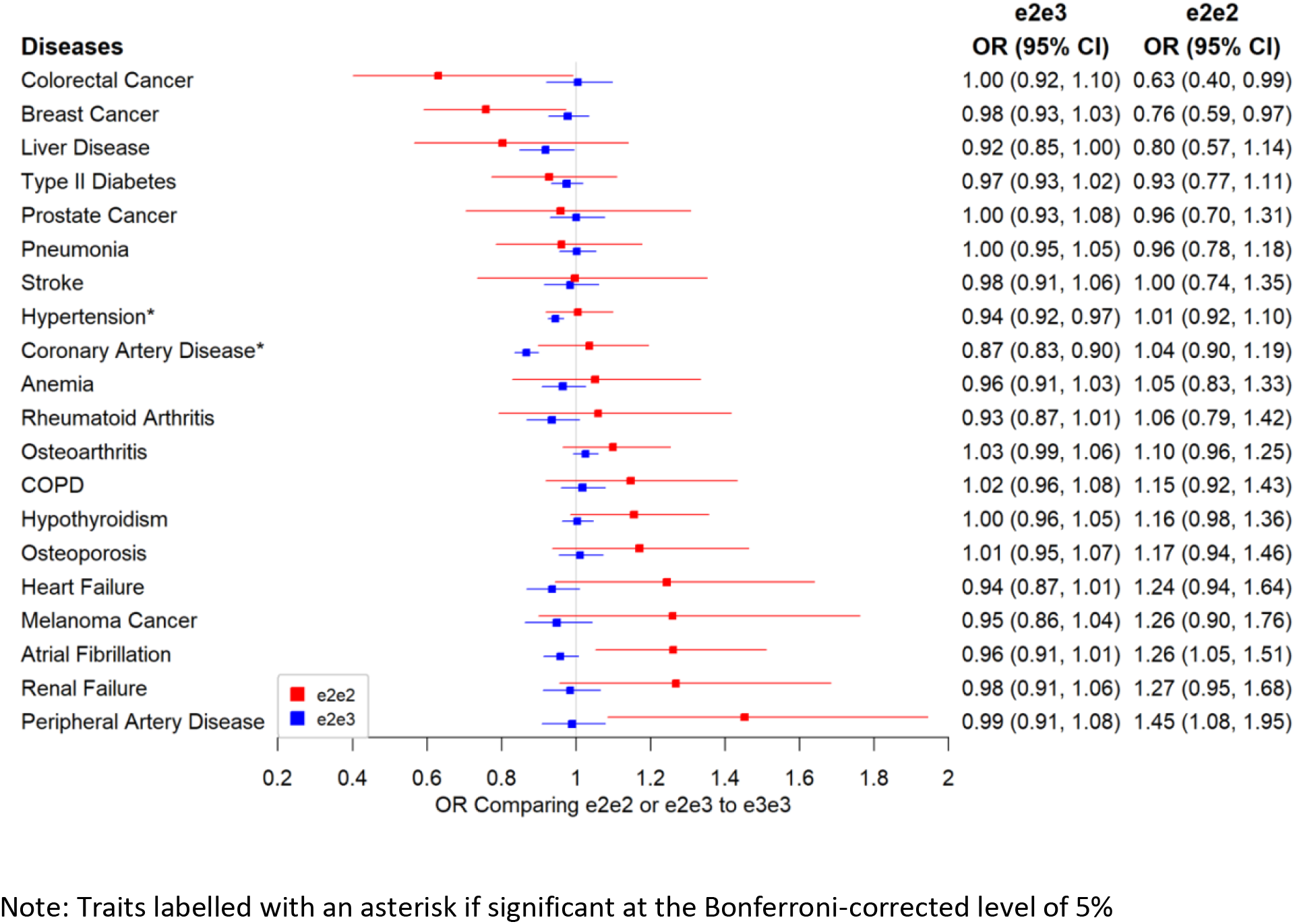
Associations Between e2 (e2e2 or e2e3) and Primary Disease Outcomes.

We found no associations between e2e3 or e2e2 and renal failure or two kidney function biomarkers, creatinine and cystatin (Supplementary Table S2). Similarly, e2e3 or e2e2 was not associated with AMD and dementia.

### Chronic Pain, Cognitive Function, Physical Measures and Mortality

e2e3 or e2e2 was not significantly associated with chronic pain, cognitive measures, and physical measures except body mass index (BMI). The mean BMI was 0.07 SD (95% CI: 0.03 to 0.11, p=0.001) higher in e2e2 than in e3e3 (Figure 4), oppositely associated with e4 (Supplementary Table S2). The hazard ratio of death during follow-up in participants comparing e2e3 to e3e3 was 0.96 (95% CI: 0.91 to 1.01, p=0.116) and that comparing e2e2 to e3e3 was 1.06 (95% CI: 0.87 to 1.30, p=0.532) (Figure 5).

**Figure 4.**
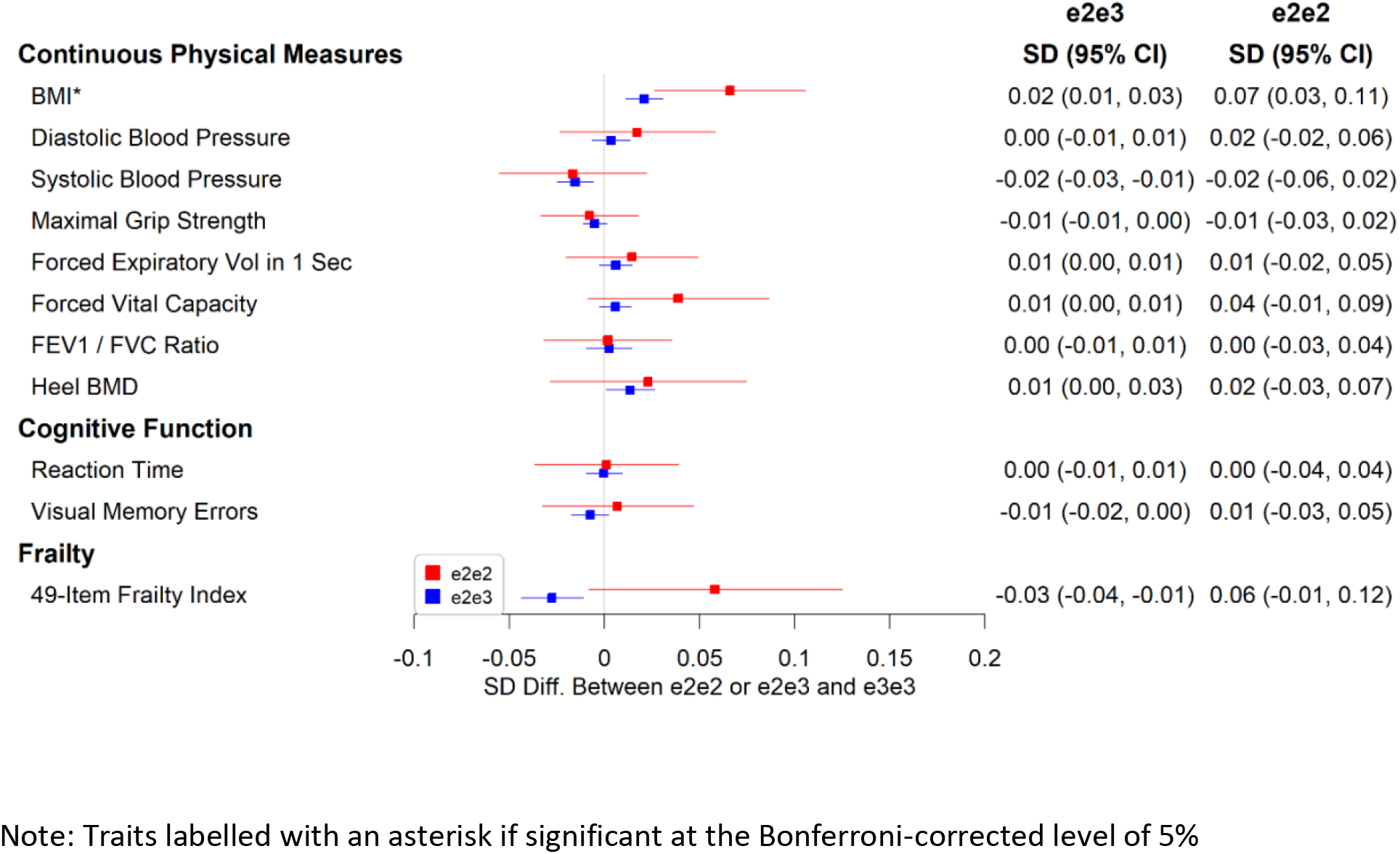
Associations Between e2e2 or e2e3 and Physical Measures, Cognitive Function, and a 49-Item Frailty.

**Figure 5.**
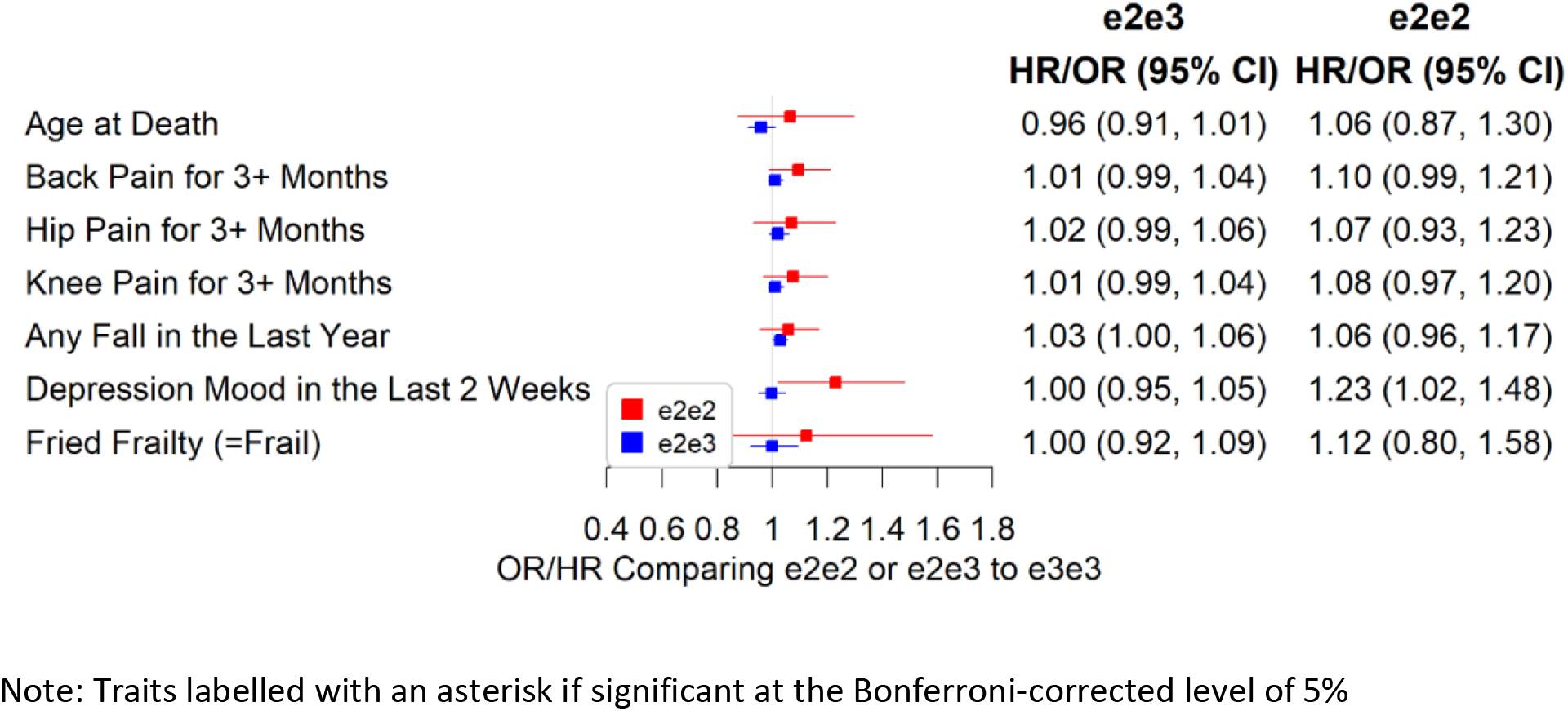
Associations Between e2e2 or e2e3 and Parent, Chronic Pain, and Physical Measures.

## Discussion

Analyses of *ApoE* e2 have reported encouraging findings of associations with longevity, in studies of parents and in centenarians [15][16]. However, there has been little data on the effects of *ApoE* e2e2 and e2e3 seperately, perhaps because the e2e2 type is relatively rare. In this large cohort analysis, we conducted a phenome-wide association study to test associations of the *ApoE* genotypes with a wide range of aging relavant traits. As *ApoE* effects could constitute potential treatment targets in aging, understanding their impacts on various aspects of aging is important, including whether e2e2 has a more powerful anti-aging effect than e2e3. We found marked reductions in total and LDL cholesterol and reductions in CAD risk in e2e3 only. However, associations for e2e2 included a marked increase in triglyceride and RDW levels and an increase in BMI (0.07 SD, 95% CI: 0.03 to 0.11, p=0.001), with no association with CAD (OR=1.04, 95% CI: 0.90 to 1.19, p=0.635). We also found minimal associations for both e2e3 and e2e2 with the studied aging outcomes, with only a small effect on Rockwood frailty (i.e., 49-item frailty) counts. The association with participant mortality in e2e3 trended in the protective direction but did not reach statistical significance, likely due to the limited follow-up thus far.

e2e2 had much lower apolipoprotein B, LDL cholesterol, total cholesterol, and lipoprotein A, and reticulocyte counts but higher apolipoprotein A1 and markedly higher triglycerides and RDW levels than e3e3. A similar pattern was found in e2e3 but the effect sizes were mostly much smaller. Several of these lipid changes are linked to CAD risk. The associations between CAD and LDL cholesterol, lipoprotein A, triglycerides, and reticulocyte count are likely to be causal based on the Mendelian randomization results [17][18][19], in which genetic variants associated with each risk factor were used to estimate associations with CAD to minimize confounding and avoid reverse causation. While the lower total and LDL-cholesterol are associated with lower CAD risk, the opposite is true for the triglyceride findings, especially the markedly higher triglyceride levels seen in e2e2, perhaps explaining the discordance in findings of e2e3 and e2e2 for CAD. The 95% confidence intervals for ORs of CAD comparing e2e2, and e2e3 to e3e3 indicate that the two ORs are quite different despite a minimal overlap: e2e3 is protective for CAD (OR=0.87, 95% CI: 0.83 to 0.90) but there was no association between e2e2 and CAD (OR=1.04, 95% CI: 0.90 to 1.19), with the point estimate trending in the opposite direction and the confidence intervals excluding a larger protective effect on CAD than seen in e2e3. Similarly, only e2e3 is protective for hypertension, but the confidence interval for the e2e2 estimate was wide and mostly right to that of e2e3, suggesting a less protective effect.

In 2012, the FDA warned that statin use may cause cognitive side effects based on post-marketing reports. However, a large randomized clinical trial of evolocumab to treat hyperlipidemia in statin users didn’t show a statistical difference in cognitive decline between the treatment and placebo groups [20]. A Mendelian randomization study also showed no evidence for a causal relationship between low LDL and dementia via genetic variation of LDL drug targets, *PCSK9* and *HMGCR* [21]. We found that e2 was strongly associated with lower LDL levels. There was no evidence suggesting that e2e2 or e2e3 was associated with dementia, but this analysis was under-powered. A recent study confirmed the protective effect of e2, substantially increased from e2e3 (OR=0.39, 95% CI: 0.30 to 0.50) to e2e2 (OR=0.13, 95% CI: 0.05 to 0.36) [22], which suggests that lower LDL is associated with reduced risk of Alzheimer’s disease through e2-related mechanisms.

The hazard ratio for participant death in this relatively young cohort was 0.96 (95% CI: 0.91 to 1.01) comparing e2e3 to e3e3, i.e., trending toward lower mortality but not reaching statistical significance. The association between e2e2 and mortality was inconclusive due to a small sample size. A meta-analysis combing four European longevity cohorts [23] showed that e2e3 was associated with increased extreme longevity (top 1% survival in the 1900 US birth cohort) (OR=1.34, 95% CI: 1.21 to 1.47) and the odds ratio comparing e2e2 to e3e3 was 1.26 (95% CI: 0.80 to 1.99). Also, the e2 determined allele of rs7412 was increasingly associated with parental extreme longevity [15], which implies the association between e2 and extreme longevity in parents as parents of participants with any e2 allele are more likely to have e2 alleles than parents of e3e3 participants. However, the association between e2e3 or e2e2 genotypes of participants and parental lifespan or longevity doesn’t imply the same association in parents. Most parents of e2e2 participants are likely e2e3 heterozygotes and several parental mating combinations can lead to e2e3 offspring. Parental lifespan and longevity outcomes therefore were not included as the main purpose of this study is to separate e2e2 and e2e3 associations with aging traits.

While associations between e2 and renal disease were previously reported, e2e3 or e2e2 was not associated with renal failure and two kidney function biomarkers, creatinine and cystatin (Supplementary Table S2). Similarly, e2e3 or e2e2 was not associated with AMD or dementia. It should be cautioned that the two conditions were rare in the UK Biobank and were under-powered to detect odds ratios ≤ 1.2 (Supplementary Table S1). With a longer follow-up, more cases may be available to retest the associations. However, we did find associations between e4 and dementia, where the ORs comparing e3e4, and e4e4 to e3e3 were 2.49 (95% CI: 2.24 to 2.78) and 7.77 (95% CI: 6.61 to 9.13), respectively.

The limitations of this study include UK Biobank selection biases, which may impact the *ApoE* genotype and aging trait association if the selection into the study is substantially associated with the aging trait [24]: such biases are likely to be modest given that ages at recruitment were 40 to 70 years old. Additionally, the presence or absence of disease was determined based on participant-reported doctor diagnoses and records during hospitalization, and the absence of primary care data in this analysis means that disease diagnoses are likely to be underestimated. Also, some participants were not old enough to develop late-onset diseases. As the sensitivity is not 100 percent, the odds ratio estimates are likely to be generally biased towards the null [25].

In conclusion, *ApoE* e2e3 was associated with reduced total and LDL cholesterol, and reduced risks of CAD and hypertension. e2e3 associations with aging measures such as frailty were modest. However, associations with e2e2 included increased triglyceride levels, increased BMI and no associations with CAD or aging measures. Overall, our results support that e2 is a potential anti-aging target but any intervention needs to take account our findings that e2e3 is likely more favorable than e2e2 for health outcomes in older groups.

## Methods

UK Biobank recruited over 500,000 participants aged 40-70 years from 2006 to 2010. A wide range of genetic and phenotypic data were collected at recruitment (baseline) and mortality and disease diagnoses were updated through linkages to death certificates, cancer registry and hospital admission records [26][27].

The DNA from blood samples was genotyped using Affymetrix UK BiLEVE Axiom array for the first ~50,000 participants and Affymetrix UK Biobank Axiom array for the rest of the cohort - the two arrays sharing over 95% marker content [27]. The two *ApoE* isoform coding SNPs, rs429358 and rs7412, on chromosome 19, were actually genotyped and the participant genotypes at these two locations were used to determine *ApoE* genotypes.

### Included Samples

To avoid genetic confounding, we analyzed European-descent participants (n=451,367, ~90% of the cohort), identified using genetic principal components analysis in detail in Thompson et al. [28]. One in third-degree or closer pairs were removed, leaving a total of 379,703, where the relatedness was determined based on pairwise kinship coefficients, calculated using genome-wide SNP data by the KING software [29].

### Aging-Related Outcomes

We classified aging-related outcomes into five categories: 1) biomarkers, 2) diseases and chronic pain, 3) mortality, 4) cognitive function, and 5) physical measures. Survival data were updated to Feb 15, 2018 and disease diagnoses to March 31, 2017. Others were surveyed or measured at recruitment/baseline.

#### Biomarkers

A panel of biomarkers from blood samples at baseline were collected including hematological measures (e.g., white blood count, red blood cell count, and hemoglobin concentration), prognostic biomarkers (e.g., lipids for vascular disease, sex hormones for cancer), diagnostic biomarkers (e.g., HbA1c for diabetes and rheumatoid factor for arthritis), and biomarkers to characterize phenotypes that are not well assessed (e.g., biomarkers for renal and liver function). The full lists including technical details can be downloaded from the links [30][31]. Each was transformed by the rank-based inverse normal transformation, followed by the z-transformation to correct distribution skewness and to unify the scale across traits.

#### Diseases and chronic pain

Disease diagnoses were either self-reported at the baseline assessment and verified by a trained nurse during the verbal interview, or from the hospital admission data (HES, hospital episode statistics, covering the period 1996 to March 31, 2017) or the cancer registry. We combined prevalent and incident cases for the analysis of *ApoE* genotype associations with likelihood of disease. A complete list of International Classification of Disease tenth revision (ICD-10) diagnosis codes used in this study is included in Supplementary Table S3.

Depression and chronic pain at baseline were assessed by survey questions to identify those with a localized pain for 3 months or longer (knee pain 3+ months, back pain 3+ months, and hip pain 3+ months) and those with depressed mood for several days or more in the past two weeks. Additionally, we derived a 49-item frailty index [32] mostly based on diseases and pains considering 60 and older only (not sensible to the middle aged), and applied log+1 transformation to correct distribution skewness.

#### Mortality

The death status was determined using the death certificate data, where age at death was calculated by date of death minus date of birth in years. For analytical purpose, we also calculated the survival time to the last follow up, which was Feb 15, 2018, for alive participants then.

#### Cognitive function

We selected two cognitive function measures from touch screen tests at baseline that covered the majority of participants, i.e., reaction time and visual memory errors. The reaction time was measured as the average time used to correctly identify a match in a symbol match game similar to the snap card game. The visual memory errors was measured as the number of errors that a participant made to complete a pairs matching task where 6 pairs of cards were presented for 3 seconds beforehand. Each was log transformed to correct skewness of the distribution. The visual memory errors was right shifted by 1 before the transformation to avoid infinite values from zero visual memory errors.

#### Physical measures

In baseline physical measures, we included body mass index (BMI), systolic and diastolic blood pressures, any falls in the last year, heel bone mineral density (BMD), lung function measures of FEV1 (forced expiratory volume in 1 second), FVC (forced vital capacity), and FEV1/FVC ratio, Fried frailty (frail or not frail), skeletal muscle mass index [33], and maximal hand grip strength. Any falls in the last year and some elements to derive the Fried frailty were assessed by survey questions. Other measurements were performed at the assessment centers when participants were recruited.

Heel bone mineral density in grams/cm^2^ was estimated based on the Quantitative Ultrasound Index through the calcaneus. The spirometry test was performed using a Vitalograph Pneumotrac 6800 that analyzed 2-3 blows of participants. The Fried frailty [34] was derived using participants aged 60 and older at baseline where the frailty status was confirmed if three or more of the conditions were met, 1) self-reported weight loss (yes/no, based on a survey question to ask weight change compared to one year ago), 2) exhaustion (yes/no, based on a survey question to ask frequency of feeling tired or having little energy over the past two weeks), 3) self-reported slow walking pace (yes/no, based on a survey question to ask usual walking pace: slow if less than 3 miles per hour), 4) lowest 20% of hand grip strength in the same sex group (yes/no), 5) lowest 20% of physical activity in the same sex group (yes/no), by the short version of International Physical Activity Questionnaire (IPAQ) [35].

The maximal hand grip strength of both hands was measured using a Jamar J00105 hydraulic hand dynamometer. The skeletal muscle mass was measured by the skeletal muscle index (SMI) defined by Janssen et al. [36],

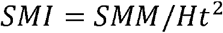

with height *(Ht)* in meters and the skeletal muscle mass *(SMM)* defined as

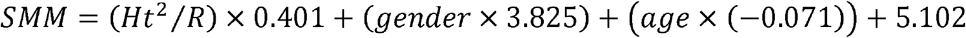

where the Bioelectrical Impedance Analysis resistance (*R*) in ohms for the whole body was taken by a Tanita BC418MA body composition analyzer; *gender* was 1 for men and 0 for women and *age* was measured in years.

## Statistical analysis

Aging-related outcomes including time-to-event (survival), continuous, and binary variables were modelled for associations with *ApoE* genotypes using Cox regression, linear regression, and logistic regression models. Prior to association analyses, continuous variables were log-transformed (cognitive function measures and 49-item frailty) or transformed by the rank-based inverse normal transformation (biomarkers) to correct distribution skewness and further z-transformed so that magnitude of mean differences by genotype between traits are comparable. Each *ApoE* genotype (e2e2, e2e3, e3e4, or e4e4) was compared with e3e3, adjusted for age at baseline (outcomes measured at baseline) or age at the last update (survival and disease outcomes), sex, assessment center, genotyping array type, and the first five genetic principal components. We highlighted associations with p-values significant at the Bonferroni-adjusted level (p<0.05/106) for the null hypothesis of no e2e3 or e2e2 effect. All the statistical analyses were performed in R version 3.4.1.

## Power analysis

To estimate power for continuous traits, we assumed that the three genotype groups e3e3, e2e3, and e2e2 share the same standard deviation for a trait. Given the sample sizes that we have for the three *ApoE* genotypes, the power to detect a 0.05 standard deviation (SD) mean difference between e3e3 and e2e3 or a 0.1 SD mean difference between e3e3 and e2e2 using an ANOVA F-test at the Bonferroni-adjusted significance level (p<0.05/106), is presented in Supplementary Table S1. Most continuous traits have over 99% power to detect the above differences, except oestradiol (65%) and rheumatoid factor (27%).

To estimate power for binary outcomes, we assumed a multiplicative e2 effect and we aimed to detect the relative risk of 1.2 comparing e2e3 to e3e3 and (1.2)^2^ comparing e2e2 to e3e3, approximately equivalent to the odds ratios from a logistic regression model as the prevalence is low (<0.1). Given the sample sizes that we have for e3e3, e2e3, and e2e2, the power to reject the null hypothesis (p<0.05/106) that both relative risks are 1, is provided in Supplementary Table S1. We focus on disease outcomes with ~80% or higher power, with melanoma cancer (79% power) and traits with better power referred to as “primary” and the rest are considered “secondary”.

Age at death was the only survival outcome in this study, with death status and age at the last follow-up information. For convenience, we approximately calculated the power using death status only as a binary outcome, which should be similar to that of the survival outcome as the death rate is low. We found in actual data analyses that the hazard ratio of death comparing e2e3 or e2e2 to e3e3 (see the Results section) was very similar to the corresponding odds ratio (results not shown).

## Data Availability

This research conducted using the UK Biobank resource, under application 14631. UK Biobank received an approval from the UK Biobank Research Ethics Committee (REC) (REC reference 11/NW/0382). All the participants provided written informed consent to participate in the study and for their data to be used in future research.

https://www.ukbiobank.ac.uk/

## Acknowledgements

This research was funded by the National Institute on Aging (R21AG060018) and conducted using the UK Biobank resource, under application 14631. UK Biobank received an approval from the UK Biobank Research Ethics Committee (REC) (REC reference 11/NW/0382). All the participants provided written informed consent to participate in the study and for their data to be used in future research.

## References

1. Suri S, Heise V, Trachtenberg AJ, Mackay CE. The forgotten APOE allele: A review of the evidence and suggested mechanisms for the protective effect of APOE e2. Neurosci Biobehav Rev [Internet]. Elsevier Ltd; 2013; 37: 2878–86. Available from: http://dx.doi.org/10.1016Zj.neubiorev.2013.10.010

2. Tudorache IF, Trusca VG, Gafencu AV. Apolipoprotein E - A Multifunctional Protein with Implications in Various Pathologies as a Result of Its Structural Features. Comput Struct Biotechnol J. The Authors; 2017; 15: 359–65.

3. Belloy ME, Napolioni V, Greicius MD. A Quarter Century of APOE and Alzheimer’s Disease: Progress to Date and the Path Forward. Neuron. 2019; 429358.

4. Eichner JE, Dunn ST, Perveen G, Thompson DM, Stewart KE. HUMAN GENOME EPIDEMIOLOGY (HuGE) REVIEW Apolipoprotein E Polymorphism and Cardiovascular Disease: A HuGE Review. Am J Epidemiol. 2002; 155: 487–95.

5. Huebbe P, Rimbach G. Evolution of human apolipoprotein E (APOE) isoforms: Gene structure, protein function and interaction with dietary factors. Ageing Res Rev [Internet]. Elsevier B.V.; 2017; 37: 146–61. Available from: http://dx.doi.Org/10.1016/j.arr.2017.06.002

6. Farrer LA, Cupples LA, Haines JL, Hyman B, Kukull WA, Mayeux R, Myers RH, Pericak-Vance MA, Risch N, Van Duijn CM. Effects of age, sex, and ethnicity on the association between apolipoprotein E genotype and Alzheimer disease: A meta-analysis. Journal of the American Medical Association. 1997. p. 1349-56.

7. Wolters FJ, Yang Q, Biggs ML, Jakobsdottir J, Li S, Evans DS, Bis JC, Harris TB, Vasan RS, Zilhao NR, Ghanbari M, Ikram MA, Launer L, et al. The impact of APOE genotype on survival: Results of 38,537 participants from six population-based cohorts (E2-CHARGE). PLoS One. 2019;.

8. Abondio P, Sazzini M, Garagnani P, Boattini A, Monti D, Franceschi C, Luiselli D, Giuliani C. The Genetic Variability of APOE in Different Human Populations and Its Implications for Longevity. Genes (Basel) [Internet]. 2019; 10: 222. Available from: https://www.mdpi.com/2073-4425/10/3/222

9. Deelen J, Evans DS, Arking DE, Tesi N, Nygaard M, Liu X, Wojczynski MK, Biggs ML, van der Spek A, Atzmon G, Ware EB, Sarnowski C, Smith A V., et al. A meta-analysis of genome-wide association studies identifies multiple longevity genes. Nat Commun. 2019; 10.

10. Klaver CC, Kliffen M, van Duijn CM, Hofman a, Cruts M, Grobbee DE, van Broeckhoven C, de Jong PT. Genetic association of apolipoprotein E with age-related macular degeneration. Am J Hum Genet. 1998; 63: 200–6.

11. Mckay GJ, Patterson CC, Chakravarthy U, Dasari S, Klaver CC, Vingerling JR, Ho L, de Jong PTVM, Fletcher AE, Young IS, Seland JH, Rahu M, Soubrane G, et al. Evidence of association of APOE with age-related macular degeneration - a pooled analysis of 15 studies. Hum Mutat. 2011; 32: 1407–16.

12. Liberopoulos E, Siamopoulos K, Elisaf M. Apolipoprotein E and Renal Disease. Am J Kidney Dis [Internet]. National Kidney Foundation, Inc.; 2004; 43: 223–33. Available from: http://dx.doi.org/10.1053/j.ajkd.2003.10.013

13. Nelson MR, Tipney H, Painter JL, Shen J, Nicoletti P, Shen Y, Floratos A, Sham PC, Li MJ, Wang J, Cardon LR, Whittaker JC, Sanseau P. The support of human genetic evidence for approved drug indications. Nat Genet [Internet]. Nature Publishing Group; 2015; 47: 856–60. Available from: http://dx.doi.org/10.1038/ng.3314

14. Kennedy BK, Berger SL, Brunet A, Campisi J, Cuervo AM, Epel ES, Franceschi C, Lithgow GJ, Morimoto RI, Pessin JE, Rando TA, Richardson A, Schadt EE, et al. Geroscience: Linking Aging to Chronic Disease. Cell [Internet]. 2014 [cited 2018 Aug 6]; 159: 709–13. Available from: http://linkinghub.elsevier.com/retrieve/pii/S009286741401366X

15. Deelen J, Evans D, Arking D, Tesi N, Nygaard M, Liu X, Wojczynski M, Biggs M, van der Spek A, Atzmon G, Ware E, Sarnowski C, Smith A, et al. A meta-analysis of genome-wide association studies identifies novel longevity genes. Nat Commun. 2019;.

16. Sebastiani P, Gurinovich A, Bae H, Andersen S, Malovini A, Atzmon G, Villa F, Kraja AT, Ben-Avraham D, Barzilai N, Puca A, Perls TT. Four Genome-Wide Association Studies Identify New Extreme Longevity Variants. J Gerontol A Biol Sci Med Sci [Internet]. 2017 [cited 2018 Aug 6]; 72: 1453–64. Available from: http://www.grg.org/adams/tables.htm

17. Holmes M V., Asselbergs FW, Palmer TM, Drenos F, Lanktree MB, Nelson CP, Dale CE, Padmanabhan S, Finan C, Swerdlow DI, Tragante V, van Iperen EPA, Sivapalaratnam S, et al. Mendelian randomization of blood lipids for coronary heart disease. Eur Heart J [Internet]. 2015; 36: 539–50. Available from: http://www.ncbi.nlm.nih.gov/pubmed/24474739

18. Burgess S, Ference BA, Staley JR, Freitag DF, Mason AM, Nielsen SF, Willeit P, Young R, Surendran P, Karthikeyan S, Bolton TR, Peters JE, Kamstrup P, et al. Association of LPA variants with risk of coronary disease and the implications for lipoprotein(a)-lowering therapies: A mendelian randomization analysis. JAMA Cardiol. 2018; 3: 619–27.

19. Astle WJ, Elding H, Jiang T, Allen D, Ruklisa D, Mann AL, Mead D, Bouman H, Riveros-Mckay F, Kostadima MA, Lambourne JJ, Sivapalaratnam S, Downes K, et al. The Allelic Landscape of Human Blood Cell Trait Variation and Links to Common Complex Disease. Cell. 2016; 167: 1415–1429.e19.

20. Calabrò P, Gragnano F, Pirro M. Cognitive function in a randomized trial of evolocumab. New England Journal of Medicine. 2017. p. 1996-7.

21. Benn M, Nordestgaard BG, Frikke-Schmidt R, Tybj^rg-Hansen A. Low LDL cholesterol, PCSK9 and HMGCR genetic variation, and risk of Alzheimer’s disease and Parkinson’s disease: Mendelian randomisation study. BMJ. 2017; 357: j1648.

22. Reiman EM, Arboleda-Velasquez JF, Quiroz YT, Huentelman MJ, Beach TG, Caselli RJ, Chen Y, Su Y, Myers AJ, Hardy J, Paul Vonsattel J, Younkin SG, Bennett DA, et al. Exceptionally low likelihood of Alzheimer’s dementia in APOE2 homozygotes from a 5,000-person neuropathological study. Nat Commun. 2020; 11.

23. Sebastiani P, Gurinovich A, Bae H, Andersen S, Malovini A, Atzmon G, Villa F, Kraja AT, Ben-Avraham D, Barzilai N, Puca A, Perls TT. Four Genome-Wide Association Studies Identify New Extreme Longevity Variants. 2017; 72: 1453–64.

24. Munafò MR, Tilling K, Taylor AE, Evans DM, Smith GD. Collider scope: When selection bias can substantially influence observed associations. Int J Epidemiol. 2018; 47: 226–35.

25. Magder LS, Hughes JP. Logistic regression when the outcome is measured with uncertainty. Am J Epidemiol. 1997; 146: 195–203.

26. Sudlow C, Gallacher J, Allen N, Beral V, Burton P, Danesh J, Downey P, Elliott P, Green J, Landray M, Liu B, Matthews P, Ong G, et al. UK Biobank: An Open Access Resource for Identifying the Causes of a Wide Range of Complex Diseases of Middle and Old Age. PLoS Med. 2015; 12.

27. Bycroft C, Freeman C, Petkova D, Band G, Elliott LT, Sharp K, Motyer A, Vukcevic D, Delaneau O, O’Connell J, Cortes A, Welsh S, Young A, et al. The UK Biobank resource with deep phenotyping and genomic data. Nature. 2018; 562: 203–9.

28. Thompson WD, Tyrrell J, Borges M-C, Beaumont RN, Knight BA, Wood AR, Ring SM, Hattersley AT, Freathy RM, Lawlor DA. Association of maternal circulating 25(OH)D and calcium with birth weight: A mendelian randomisation analysis. PLoS Med [Internet]. 2019; 16: e1002828. Available from: http://www.ncbi.nlm.nih.gov/pubmed/31211782

29. Manichaikul A, Mychaleckyj JC, Rich SS, Daly K, Sale M, Chen WM. Robust relationship inference in genome-wide association studies. Bioinformatics. 2010; 26: 2867–73.

30. Sheard SM, Nicholls R, Froggatt J. UK Biobank Haematology Data Companion Document [Internet]. 2017. Available from: https://biobank.ctsu.ox.ac.uk/crystal/crystal/docs/haematology.pdf

31. UK Biobank biomarker panel [Internet]. [cited 2019 Oct 28]. Available from: http://www.ukbiobank.ac.uk/wp-content/uploads/2013/12/ukb_biomarker_panel_final_website_Oct2013_CLMS.pdf

32. Williams DM, Jylhava J, Pedersen NL, Hagg S. A frailty index for UK Biobank participants. Journals Gerontol Med Sci. 2018;.

33. Janssen I, Heymsfield SB, Baumgartner RN, Ross R. Estimation of skeletal muscle mass by bioelectrical impedance analysis. J Appl Physiol. 2000; 89: 465–71.

34. Fried LP, Tangen CM, Walston J, et al, Newman AB, Hirsch C, Gottdiener J, Seeman T, Tracy R, Kop WJ, Burke G, McBurnie MA, et al, et al. Frailty in Older Adults: Evidence for a Phenotype. Journals Gerontol Ser A Biol Sci Med Sci [Internet]. 2001; 56: M146- 57. Available from: http://www.ncbi.nlm.nih.gov/pubmed/11253156%0Ahttps://academic.oup.com/biomedgerontology/article-lookup/doi/10.1093/gerona/56.3.M146

35. Craig CL, Marshall AL, Sjostrom M, Bauman AE, Booth ML, Ainsworth BE, Pratt M, Ekelund U, Yngve A, Sallis JF, Oja P. International physical activity questionnaire: 12-country reliability and validity. Med Sci Sports Exerc. Canadian Fitness and Lifestyle Research Institute, Ottawa, Canada.; 2003; 35: 1381–95.

36. Janssen I, Heymsfield SB, Baumgartner RN, Ross R. Estimation of skeletal muscle mass by bioelectrical impedance analysis. J Appl Physiol. 2017; 89: 465–71.

